# Uncertainty-Gated Glaucoma Screening: Combining Semi-Supervised Classification with Multi-Agent Large Language Model Deliberation

**DOI:** 10.64898/2026.04.17.26351127

**Authors:** Sai Varun Garimella Narasimha, Nicholas Brown, Srinivas Sridhar

**Author notes:** **Correspondence to:** Sai Varun Garimella Narasimha.

## Abstract

Automated glaucoma screening from optical coherence tomography (OCT) faces two persistent challenges: scarcity of expert-labeled data and unreliable model predictions on diagnostically ambiguous cases. We present a two-tier diagnostic pipeline that addresses both. In the first tier, an EfficientNetV2-S classifier trained under a semi-supervised pseudo supervisor framework achieves 0.84 AUC on 150 held-out test patients from the Harvard Glaucoma Detection and Progression dataset, using only 350 labeled training samples out of 700. In the second tier, 124 flagged cases are routed to a multi-agent system built on MedGemma 4B, where three specialist agents deliberate over three rounds before rendering a final diagnosis. On these flagged cases, the agent system achieves 100% sensitivity—detecting all 55 glaucoma cases with zero missed diagnoses—and 89.5% overall accuracy (111/124), compared to the classifier’s 73.4% (91/124). Uncertainty analysis confirms that the classifier’s output probability reliably separates confident predictions (96.3% accuracy, *n* = 27) from uncertain ones (74.0%, *n* = 123), producing a 22-percentage-point gap that serves as a triage signal. The agents fix 32 cases the classifier misclassifies while introducing 12 new errors, yielding a net improvement of 20 cases. These results are from a single training run without variance estimates and should be interpreted as preliminary evidence that uncertainty-gated routing to vision-language model agents can meaningfully improve diagnostic accuracy on the cases where automated classifiers are least reliable.

## 1 Introduction

Glaucoma is the leading cause of irreversible blindness worldwide, affecting an estimated 80 million people with projections exceeding 111 million by 2040 [Tham et al., 2014]. The disease damages retinal ganglion cells and their axons in the retinal nerve fiber layer (RNFL), producing characteristic structural thinning detectable by optical coherence tomography (OCT) and functional visual field loss measurable by automated perimetry. Early detection is critical because glaucomatous damage is irreversible, yet the disease is often asymptomatic until advanced stages. Automated screening systems based on deep learning have shown promise for glaucoma detection from OCT imaging. However, two challenges limit their clinical deployment. First, obtaining expert-labeled ophthalmic data is expensive and time-consuming, creating a persistent label scarcity problem. Second, classifiers produce unreliable predictions on diagnostically ambiguous cases—patients whose features fall near the decision boundary—yet these are precisely the cases where clinical consequences of misclassification are most severe.

Semi-supervised learning offers a partial solution to label scarcity. The pseudo supervisor framework [Luo et al., 2023] uses reinforcement learning to train a secondary network that generates pseudo-labels for unlabeled data, with the reward signal derived from whether those labels improve the primary classifier’s validation performance. This approach has demonstrated competitive performance on the Harvard Glaucoma Detection and Progression (Harvard-GDP) dataset even when a substantial fraction of training labels are withheld.

The second challenge—unreliable predictions on ambiguous cases—is not resolved by uncertainty quantification alone. Methods such as MC dropout [Gal and Ghahramani, 2016] and deep ensembles [Lakshminarayanan et al., 2017] can identify *which* cases are uncertain, but they do not resolve the uncertainty. A diagnostically ambiguous case flagged as uncertain still requires a mechanism capable of deeper reasoning—one that can integrate multiple data modalities, apply domain-specific clinical knowledge, and weigh structural and functional evidence in the way a specialist would. This motivates a secondary evaluation system that goes beyond confidence estimation to perform multimodal clinical reasoning on the flagged cases.

Recent advances in medical vision-language models (VLMs) provide a mechanism for this secondary evaluation. Models such as MedGemma [Sellergren et al., 2025] can jointly process medical images and clinical text, enabling structured reasoning that integrates structural imaging findings with functional test results and demographic context. Research on multi-agent deliberation has further shown that multiple language model instances adopting distinct specialist roles and engaging in structured discussion can improve diagnostic performance on complex clinical tasks [Wang, 2025].

Most existing automated glaucoma screening systems operate on a single data modality—typically OCT structural imaging alone—and produce a single prediction without distinguishing between cases where the model is confident and cases where it is not. This creates two sources of diagnostic error. First, structural imaging alone cannot capture functional vision loss, meaning patients with early or atypical glaucoma that manifests primarily in visual field deficits may be missed. Second, when a single-modality classifier encounters an ambiguous case near its decision boundary, it has no mechanism to seek additional evidence—it must commit to a diagnosis with whatever limited information it has. In clinical practice, an ophthalmologist encountering an ambiguous structural finding would order a visual field test and correlate the two; current automated systems lack this adaptive behavior. Our approach addresses both sources of error. By routing uncertain cases to an agent system that integrates OCT and visual field data, we bring in a complementary diagnostic modality precisely where it is most needed—on the cases the classifier struggles with. Furthermore, the multi-agent design functions as a form of diagnostic ensemble: rather than relying on a single model’s interpretation, three specialist agents independently assess the evidence from different clinical perspectives and refine their positions through structured deliberation before a director synthesizes all opinions. This ensemble-style reasoning reduces the risk of any single misinterpretation driving the final diagnosis, which is particularly important for sensitivity—catching every glaucoma case—where the cost of a single missed diagnosis is irreversible vision loss.

In this work, we implement this approach as an uncertainty-gated two-tier pipeline. The first tier is a semi-supervised EfficientNetV2-S [Tan and Le, 2021] classifier that screens all patients and identifies cases where its prediction confidence falls below a defined threshold. The second tier is a multi-agent system in which three MedGemma 4B agents—an OCT structural specialist, a visual field functional specialist, and a clinical director—evaluate flagged cases through three rounds of cross-specialty deliberation before rendering a final diagnosis. Critically, the agent system has access to both OCT imaging and visual field data, whereas the classifier operates on OCT alone, providing the agents with an additional data modality unavailable to the first tier. We evaluate this pipeline on the Harvard-GDP dataset [Luo et al., 2023], a publicly available collection of 1,000 patients with paired RNFL thickness maps, visual field total deviation scores, and demographic information. Our contributions are: (1) demonstrating that classifier output probability reliably separates confident from uncertain predictions, producing a triage signal with a 22-percentage-point accuracy gap; and (2) designing and evaluating a three-agent, three-round deliberation protocol that achieves 100% sensitivity on flagged glaucoma cases while improving overall accuracy on those cases by 16.1 percentage points. Figure 1 illustrates the complete pipeline architecture.

**Figure 1:**
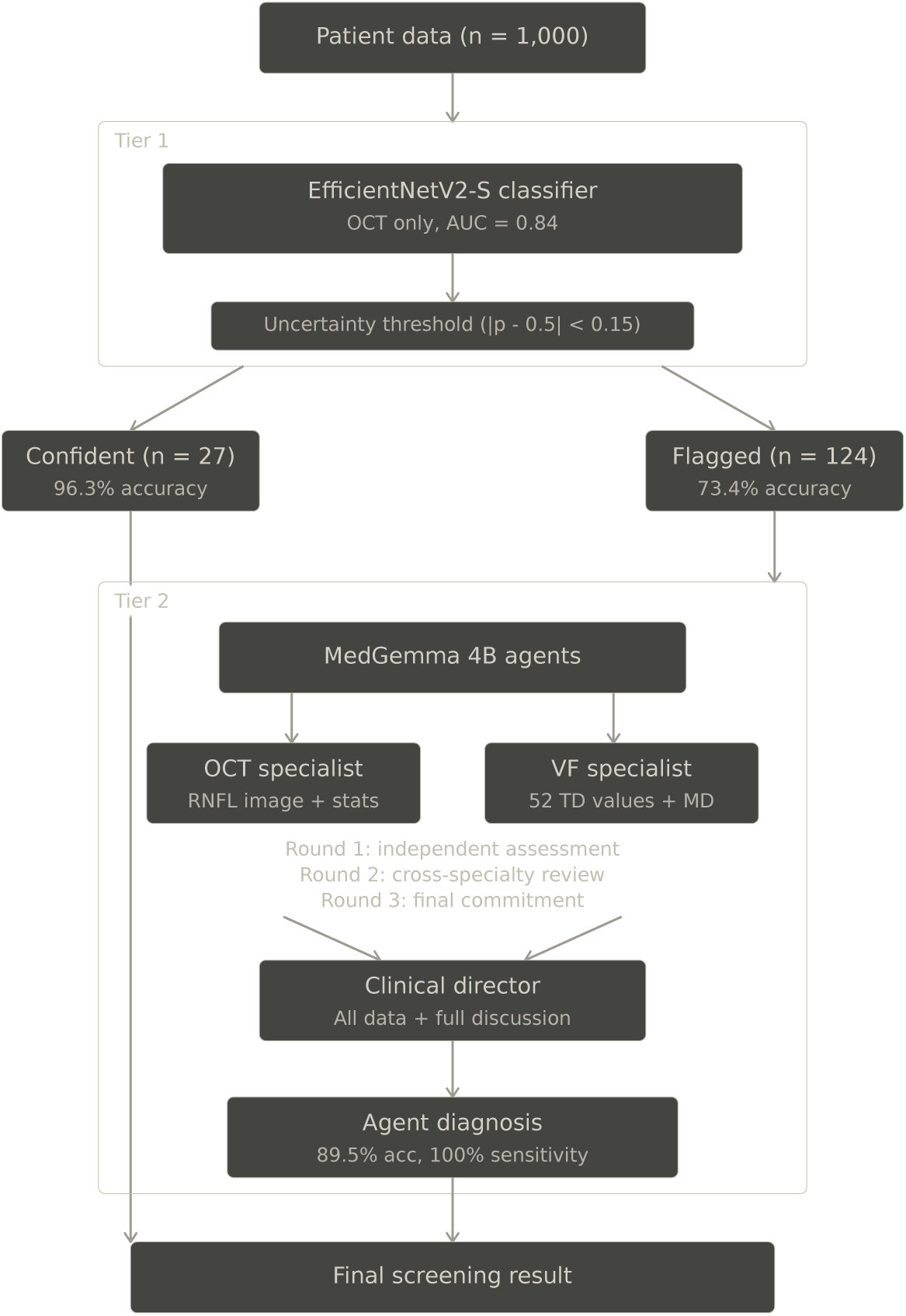
Uncertainty-gated two-tier screening pipeline. Tier 1: an EfficientNetV2-S classifier screens all patients using OCT data alone. Uncertainty analysis separates confident predictions (*n* = 27, 96.3% accuracy) from flagged cases (*n* = 124, 73.4% accuracy). Tier 2: flagged cases are evaluated by a multi-agent system (MedGemma 4B) comprising an OCT structural specialist, a visual field functional specialist, and a clinical director, deliberating over three rounds. The agent system achieves 89.5% accuracy with 100% sensitivity on the flagged cases.

## 2 Methods

### 2.1 Dataset

We use the Harvard Glaucoma Detection and Progression (Harvard-GDP) dataset [Luo et al., 2023], published at the IEEE/CVF International Conference on Computer Vision in 2023. The dataset comprises 1,000 patients stored as individual NumPy compressed files (data_0001.npz through data_1000.npz). Each patient record contains: a 225 × 225 pixel RNFL thickness map representing a 6×6 mm peripapillary scan centered on the optic nerve head; 52 total deviation (TD) scores from Humphrey 24-2 visual field testing; a mean deviation (MD) value summarizing overall visual field performance; and demographic variables including age, gender, race, and Hispanic ethnicity.

#### 2.1.1 Labeling Convention

Following the convention established in the Harvard-GDP paper, we derive binary labels from the MD value: patients with MD ≥ −1 dB are labeled healthy (label = 1) and patients with MD *<* −1 dB are labeled glaucoma (label = 0). This threshold-based labeling from a continuous variable introduces a hard boundary at −1 dB; patients near this threshold may be clinically ambiguous regardless of their assigned label.

#### 2.1.2 Data Partitioning

The 1,000 patient files were shuffled with a fixed random seed (seed = 42) and partitioned into 700 training, 150 validation, and 150 test patients. Table 1 reports the class distribution across splits.

**Table 1:** Class distribution across data splits.

| Split | Glaucoma ($MD < -1$) | Healthy ($MD \geq -1$) | Total |
| --- | --- | --- | --- |
| Training | 306 (43.7%) | 394 (56.3%) | 700 |
| Validation | 65 (43.3%) | 85 (56.7%) | 150 |
| Test | 71 (47.3%) | 79 (52.7%) | 150 |
| Total | 442 (44.2%) | 558 (55.8%) | 1,000 |

#### 2.1.3 Semi-Supervised Label Masking

To simulate clinical label scarcity, 50% of training labels (350 patients) were randomly masked by setting their labels to −100, a sentinel value that instructs the training loop to treat these samples as unlabeled. The remaining 350 training patients retain their labels. Validation and test sets are fully labeled.

### 2.2 Image Preprocessing

Each 225 × 225 RNFL thickness map is preprocessed as follows: (1) the first row and first column are removed, yielding a 224 × 224 image compatible with EfficientNetV2-S input requirements; (2) pixel values are clipped to the range [−2, 350] and shifted by adding 2, producing a non-negative range of [0, 352]; (3) the array is converted to float32 and reshaped to a single-channel tensor of dimensions (1, 224, 224).

### 2.3 Classification Model

#### 2.3.1 Architecture

The classifier is an EfficientNetV2-S [Tan and Le, 2021] pretrained on ImageNet. Two modifications adapt it for single-channel OCT input and binary classification: the first convolutional layer is replaced with a Conv2d(1, 24, 3 × 3, stride = 2, padding = 1) layer to accept one input channel instead of three, and the final classification head is replaced with a Linear(1280, 1) layer followed by a sigmoid activation. The model contains 20,178,337 trainable parameters.

#### 2.3.2 Pseudo Supervisor Framework

We train the classifier under the generalization-reinforced pseudo-supervised learning (GRSL) framework described by Luo et al. [2023]. A second network, the pseudo supervisor, shares the same EfficientNetV2-S architecture and learns to generate pseudo-labels for the 350 unlabeled training samples.

At each training step, the pseudo supervisor produces a probability for each unlabeled image, from which a binary pseudo-label is sampled via a Bernoulli distribution. The classifier’s validation loss is measured before and after training on the pseudo-labeled batch. The difference determines the reward:

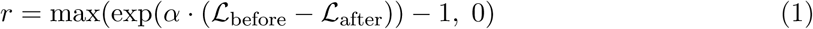

where L_before_ and L_after_ are binary cross-entropy losses on a validation mini-batch evaluated before and after the classifier update, and *α* = 1.0 controls reward sensitivity. Positive reward indicates the pseudo-labels improved the classifier; zero reward indicates they did not.

Every 50 steps (*β* = 50), accumulated rewards are discounted with factor *γ* = 0.9 and normalized by subtracting the mean and dividing by the standard deviation. The pseudo supervisor is then updated via the REINFORCE policy gradient:

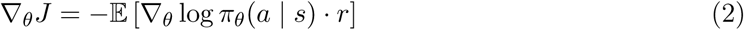

where *π_θ_* is the pseudo supervisor’s policy, *s* is the unlabeled image, *a* is the sampled pseudo-label, and *r* is the normalized reward.

The validation mini-batches used for reward computation are drawn from the 150-patient validation set through an infinite cycling iterator with batch size 16. This validation set is the same one used for epoch-level model selection (Section 2.3.4), meaning the reward signal and the selection criterion share the same data pool. We discuss the implications of this overlap in Section 4.5.

#### 2.3.3 Training Configuration

Both networks are optimized with AdamW (weight decay = 0). The classifier uses a learning rate of 4 × 10*^−^*^5^ and the pseudo supervisor uses 2 × 10*^−^*^5^. Training proceeds for 20 epochs with batch size 8. The loss function is binary cross-entropy. Training was conducted on a single NVIDIA T4 GPU in Google Colab.

#### 2.3.4 Model Selection

At the end of each epoch, the classifier is evaluated on the full validation set. The model checkpoint with the highest validation AUC is saved. Test set metrics are computed once on this saved checkpoint and are reported below. The test set was not used for model selection or any training decision.

### 2.4 Uncertainty Analysis

The classifier outputs a probability *p* ∈ [0, 1] for each test patient, where values near 1.0 indicate confidence toward healthy and values near 0.0 indicate confidence toward glaucoma. We define an uncertainty zone as |*p* − 0.5| *< δ*, corresponding to predicted probabilities between 0.5 − *δ* and 0.5 + *δ*. We set *δ* = 0.15 as a fixed design parameter prior to examining test predictions, yielding an uncertainty range of [0.35, 0.65].

Patients are categorized into three groups: *confident correct* (prediction outside the uncertainty zone and matching the true label), *confident wrong* (prediction outside the uncertainty zone but not matching the true label), and *uncertain* (prediction within the uncertainty zone). All uncertain patients plus any confident-wrong patients are flagged for agent review.

### 2.5 Multi-Agent Diagnostic System

Figure 1 provides an overview of the complete two-tier pipeline before we describe the agent system in detail.

#### 2.5.1 Model

The agent system uses MedGemma 4B [Sellergren et al., 2025], a medical vision-language model from Google capable of processing both medical images and clinical text. The model is loaded in bfloat16 precision with automatic device mapping. Generation uses greedy decoding (do_sample=False) with a maximum of 300 new tokens per response.

#### 2.5.2 Agent Roles

Three specialist agents are instantiated, each with a distinct clinical persona and data access:

##### OCT Structural Specialist (“Dr. Chen”)

Receives the RNFL thickness map as a PIL image and three computed statistics: average thickness, maximum thickness, and standard deviation. Minimum thickness was excluded because it equals −2.0 for all patients in the dataset—a preprocessing artifact, not a clinical measurement. The prompt embeds clinical knowledge about the ISNT rule (RNFL is thickest inferiorly, then superiorly, nasally, temporally), characteristic glaucomatous damage patterns (inferior and superior bundle loss, wedge-shaped defects, asymmetry), and a critical correction: the computed map average is lower than standard clinical peripapillary norms (∼97 *µ*m) because the map includes background pixels outside retinal tissue.

##### Visual Field Functional Specialist (“Dr. Patel”)

Receives all 52 TD values as text and summary statistics: MD, average deviation, worst point, best point, count of points below −10 dB, and count of points in the normal range (−5 to 5 dB). The prompt embeds knowledge about MD severity categories, glaucomatous visual field patterns (nasal steps, arcuate scotomas, paracentral scotomas), and the clinical significance of clustered versus scattered depressions. This agent operates in text-only mode without image input.

##### Clinical Director (“Dr. Williams”)

Receives all data available to both specialists—the RNFL image, all 52 TD values, computed statistics, and demographic information—plus the full three-round discussion transcript and the classifier’s prediction with its confidence. The prompt explicitly states that the classifier prediction is unreliable and instructs the director to make an independent clinical judgment.

#### 2.5.3 Deliberation Protocol

Each flagged patient undergoes three rounds of deliberation:

##### Round 1 (Independent Assessment)

Each specialist evaluates the patient independently using a four-step chain-of-thought prompt: (1) examine the primary data source; (2) identify damage patterns; (3) consider summary statistics in context; (4) state a concern level (None, Mild, Moderate, or Severe).

##### Round 2 (Cross-Specialty Review)

Each specialist receives their own Round 1 assessment alongside the other specialist’s Round 1 assessment, with instructions to update their opinion in light of cross-modal evidence. The OCT specialist is reminded that structural damage can precede functional loss; the VF specialist is reminded that structural findings increase the clinical significance of mild functional depressions.

##### Round 3 (Final Commitment)

Each specialist receives the full discussion history (both Round 1 and Round 2 responses) and is required to commit to a final position using one of two exact phrases: “My final assessment: this patient has glaucoma” or “My final assessment: this patient is healthy.”

##### Director Decision

The director receives all six specialist responses, the raw data, the RNFL image, demographic information, and the classifier’s output. A five-step chain-of-thought prompt guides the decision: (1) independently examine the RNFL image; (2) independently review the 52 VF values; (3) assess structure-function agreement; (4) consider demographic risk factors (age, race); (5) weigh team opinions. The director outputs a final diagnosis (Glaucoma or Healthy), a confidence level (High, Medium, or Low), and 2–3 sentences of clinical reasoning.

#### 2.5.4 Prompt Design Principles

Three design choices informed by the literature on medical LLM prompting were applied across all agent prompts. First, named expert personas with quantified experience (e.g., “15 years of experience, reviewed over 20,000 RNFL thickness maps”) were used rather than generic role descriptions. Second, domain-specific clinical knowledge was embedded directly in the prompt context rather than relying on the model’s parametric knowledge. Third, chain-of-thought reasoning with numbered steps was imposed to encourage systematic evaluation before diagnosis [Wei et al., 2022, Savage et al., 2024]. Complete prompt texts for all agents and rounds are provided in Appendix A.

#### 2.5.5 Diagnosis Parsing

The director’s free-text response is parsed to extract a binary diagnosis. The parser first searches for a “Final Diagnosis” line and checks whether it contains “Glaucoma” or “Healthy.” If no such line is found, it falls back to scanning the last 200 characters of the response, then the full response. Responses that cannot be parsed are flagged as “UNPARSED” and excluded from accuracy calculations. In the reported evaluation, all 124 responses were successfully parsed.

## 3 Results

### 3.1 Classifier Training

Training loss decreased from 0.6511 at epoch 1 to 0.3240 at epoch 20. The best validation AUC occurred at epoch 1 (val AUC = 0.8599), with no subsequent epoch exceeding this value (Figure 2). Test AUC at the selected epoch was 0.8408.

**Figure 2:**
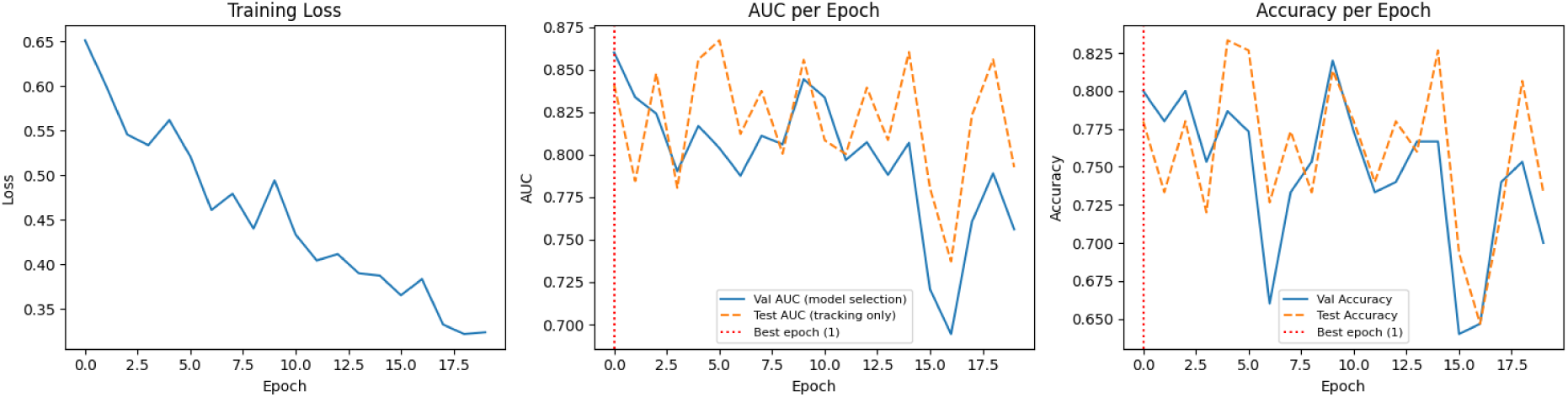
Training progress across 20 epochs. **Left:** Training loss decreases steadily. **Center:** Validation AUC (solid blue, used for model selection) peaks at epoch 1 (red dashed line); test AUC (dashed orange) is tracked but not used for selection. **Right:** Validation and test accuracy.

### 3.2 Classifier Test Performance

The epoch-1 model was evaluated once on the 150-patient test set. Table 2 reports the results.

**Table 2:** Classifier performance on the 150-patient test set (epoch-1 model selected by validation AUC).

| Metric | Value |
| --- | --- |
| AUC | 0.8408 |
| Accuracy | 0.7800 |
| F1 Score (macro) | 0.7800 |
| Sensitivity (glaucoma detection) | 0.8169 (58/71) |
| Specificity (healthy identification) | 0.7468 (59/79) |
| Missed glaucoma (false negatives) | 13 |
| False alarms (false positives) | 20 |

Figure 3 shows the distribution of predicted probabilities and the ROC curve.

**Figure 3:**
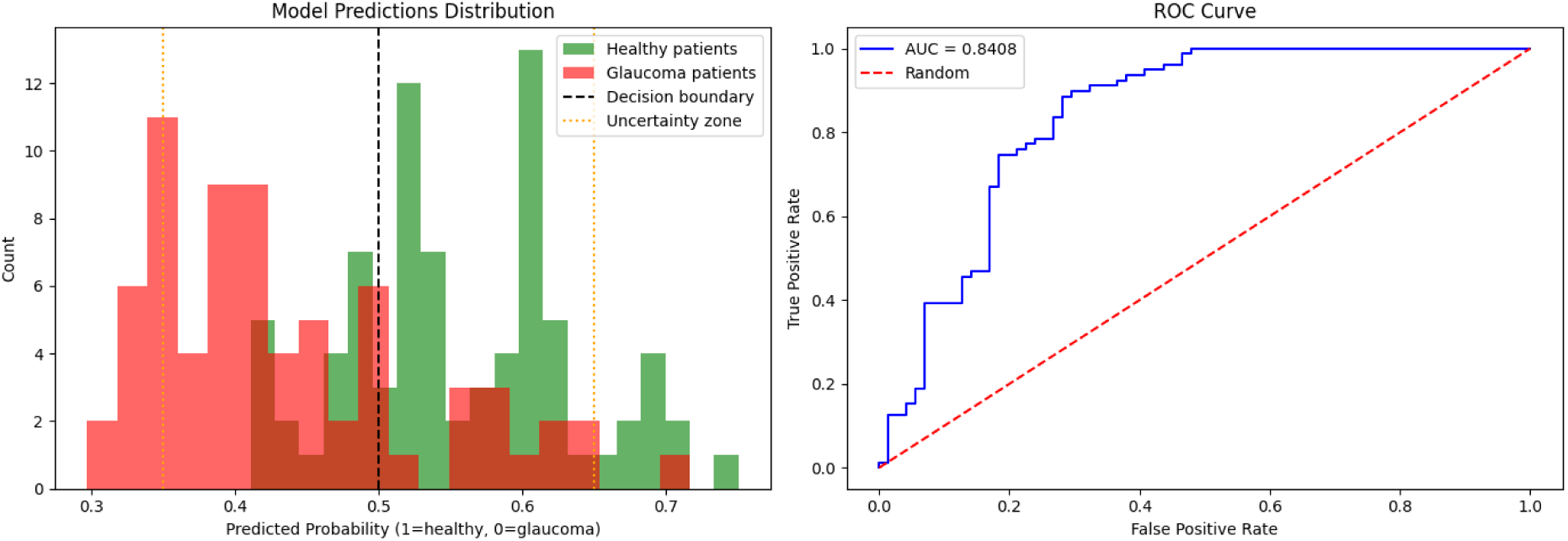
Left: Distribution of classifier predicted probabilities for healthy (green) and glaucoma (red) patients. The decision boundary at 0.5 (black dashed) and the uncertainty zone boundaries at 0.35 and 0.65 (orange dotted) are indicated. **Right:** ROC curve with AUC = 0.8408.

For reference, Luo et al. [2023] reported AUC values of 0.8693 (supervised baseline), 0.8727 (pseudo supervisor), and 0.8818 (pseudo supervisor with augmentation) on their full training set of 15,725 samples. Our lower AUC is expected given the substantially smaller training set (350 labeled samples versus 15,725) and the absence of data augmentation.

### 3.3 Uncertainty Analysis

Table 3 presents the uncertainty stratification of the 150 test patients.

**Table 3:**
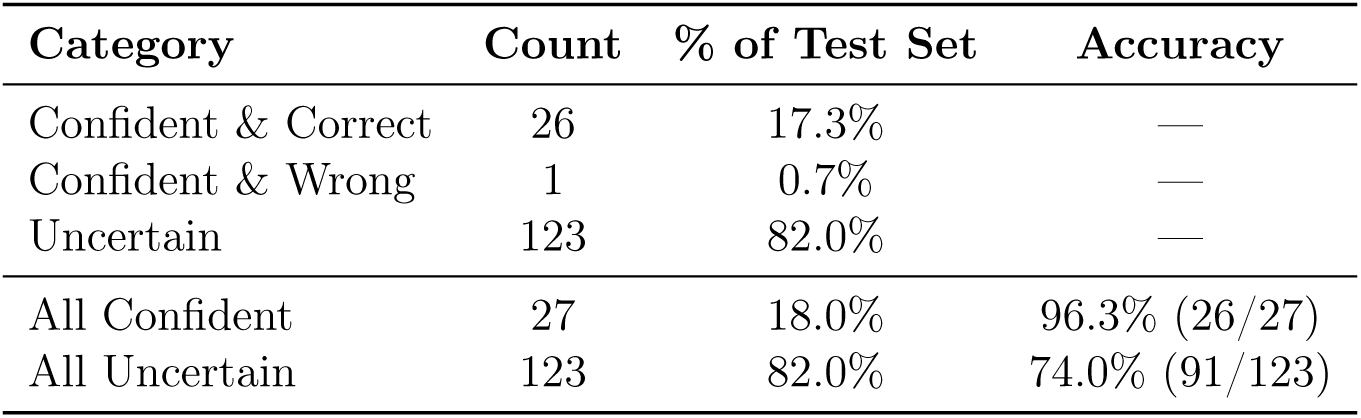
Uncertainty stratification of test predictions (*δ* = 0.15, set prior to examining test data).

| Category | Count | % of Test Set | Accuracy |
| --- | --- | --- | --- |
| Confident & Correct | 26 | 17.3% | — |
| Confident & Wrong | 1 | 0.7% | — |
| Uncertain | 123 | 82.0% | — |
| All Confident | 27 | 18.0% | 96.3% (26/27) |
| All Uncertain | 123 | 82.0% | 74.0% (91/123) |

The 22-percentage-point accuracy gap between confident predictions (96.3%) and uncertain predictions (74.0%) confirms that the classifier’s output probability functions as a triage signal. We note that 96.3% is based on only 27 patients, and this small sample size limits the precision of this estimate. The high proportion of uncertain cases (82.0%) reflects the model’s limited confidence, likely a consequence of the epoch-1 checkpoint representing minimal fine-tuning beyond the pretrained features.

A total of 124 patients were flagged for agent review: 123 uncertain cases plus 1 confident misclassification.

### 3.4 Agent System Performance

All 124 flagged patients were processed through the three-round deliberation protocol. Processing averaged approximately 85 seconds per patient (total runtime ≈ 3 hours on a single T4 GPU). All director responses were successfully parsed. Table 4 presents the head-to-head comparison.

**Table 4:** Classifier versus agent system performance on the 124 flagged cases.

| Metric | Classifier | Agent System |
| --- | --- | --- |
| Overall Accuracy | 73.4% (91/124) | 89.5% (111/124) |
| *Glaucoma patients (n = 55)* |  |  |
| Correctly detected | 76.4% (42/55) | 100.0% (55/55) |
| Missed | 13 | 0 |
| *Healthy patients (n = 69)* |  |  |
| Correctly cleared | 71.0% (49/69) | 81.2% (56/69) |
| False alarms | 20 | 13 |
| Sensitivity | 0.7636 | 1.0000 |
| Specificity | 0.7101 | 0.8116 |
| Cases fixed (classifier wrong → agent right) | — | 32 |
| Cases broken (classifier right → agent wrong) | — | 12 |
| Net improvement | — | +20 |

The most clinically significant result is 100% sensitivity: all 55 glaucoma patients among the flagged cases were correctly identified, with zero missed diagnoses. We note that the one-sided 97.5% confidence interval for 55/55 observations has a lower bound of approximately 93.5%, so the true sensitivity may be lower than 100% in a larger sample. The 13 agent errors are all false positives (healthy patients diagnosed as glaucoma); the agent produces zero false negatives on this subset.

### 3.5 Combined System Performance

The two-tier system partitions the 150 test patients into two groups handled by different mechanisms. The 27 confident cases are handled by the classifier alone at 96.3% accuracy. The 124 flagged cases are handled by the agent system at 89.5% accuracy.

## 4 Discussion

### 4.1 Uncertainty as a Triage Signal

The 22-percentage-point gap between confident and uncertain accuracy suggests that classifier output probability, without any additional calibration, is a potentially useful triage signal. Confident predictions can be trusted; uncertain predictions require further evaluation. This finding aligns with prior work on uncertainty quantification in deep learning [Gal and Ghahramani, 2016, Lakshminarayanan et al., 2017], which has established that predictive uncertainty correlates with error likelihood.

The high proportion of uncertain cases (82%) warrants discussion. This is substantially higher than would be expected in a well-calibrated clinical system and likely reflects two factors: the epoch-1 checkpoint has undergone minimal fine-tuning beyond ImageNet pretraining, limiting its discriminative confidence on domain-specific features; and the 350-label training regime provides insufficient data for a 20-million-parameter model to develop sharp decision boundaries. The pattern of declining training loss with non-improving validation performance from the first epoch is consistent with overfitting, suggesting that the pretrained ImageNet features provide the best generalization achievable at this data scale.

An important question raised by the epoch-1 peak is whether the pseudo supervisor framework contributes anything beyond the pretrained ImageNet features at this data scale. A comparison to a frozen-feature baseline (ImageNet features with a linear probe) would clarify this and is planned for future work.

### 4.2 Agent System Strengths and Confounds

The agent system’s 100% sensitivity on flagged glaucoma cases is the most clinically relevant result. In glaucoma screening, missed diagnoses carry asymmetric consequences—undetected disease leads to irreversible vision loss, while false positives lead to additional testing. The agent system’s error profile (13 false positives, 0 false negatives on flagged cases) aligns with the clinical priority of maximizing detection.

However, three confounds are entangled in the agent system’s improvement, and we cannot attribute the gains to any single design choice. First, and most importantly, the agents receive visual field data that the classifier never sees—an independently diagnostic modality. Providing any system with a second data modality that is independently informative about the outcome will improve performance, regardless of architecture [Xiong et al., 2022]. Second, the agents use a different model (MedGemma 4B versus EfficientNetV2-S). Third, the agents use a multi-round deliberation protocol. Without ablations—a single MedGemma call with both modalities, multi-agent without deliberation rounds, or the classifier retrained with visual field features—it is impossible to isolate the contribution of multi-agent deliberation from the contribution of additional data. The improvement may be driven primarily by the additional modality rather than the architectural novelty. Future work should include these ablations.

The three-round deliberation protocol allows specialists to update their assessments in light of cross-modal evidence, enabling structure-function correlation that neither specialist could perform alone. The chain-of-thought prompting forces systematic evaluation rather than pattern-matching. Whether these specific design choices contribute beyond simply providing both modalities to a single model remains an open empirical question.

### 4.3 Computational Cost

The agent system processes each patient in approximately 85 seconds on a single T4 GPU, requiring 7 sequential LLM calls per patient (two per round for specialists, plus the director). For the 124 flagged patients, total runtime was approximately 3 hours. In a clinical screening deployment processing thousands of patients, this computational cost would be a significant consideration. The two-tier design mitigates this: only uncertain cases (here, 82% of the test set, but potentially a smaller fraction with a better-calibrated first-tier classifier) require the expensive agent evaluation. Reducing the proportion of uncertain cases through data augmentation or improved training is a priority for practical deployment.

### 4.4 Prompt Sensitivity

The current prompts represent the fifth iteration of the agent prompt design. Earlier versions exhibited dramatic performance swings: one version achieved only 18.2% sensitivity (agents classified nearly all patients as healthy), while another over-diagnosed glaucoma. The key modifications that stabilized performance were embedding the ISNT rule, correcting for background pixel effects on average thickness, providing MD severity categories, and removing the minimum thickness artifact. This sensitivity to prompt design is itself a significant finding, suggesting that MedGemma 4B’s diagnostic accuracy is heavily dependent on the quality of contextual guidance rather than its parametric medical knowledge alone.

### 4.5 Limitations

#### Single training run

All results are from one training run with one random seed. GPU-level non-determinism and random data splits mean the exact numbers will vary across runs. Confidence intervals and multi-seed runs are planned for future work.

#### Shared validation set

The validation set serves dual purposes: computing the RL reward signal during training (via mini-batch sampling) and selecting the best model checkpoint (via full-set evaluation). This overlap means the model selection criterion is not fully independent of the training signal. The reward signal shapes the training trajectory of the pseudo supervisor, and the same data then determines which checkpoint is saved. This coupling may introduce optimistic bias in the reported validation AUC and, by extension, in the quality of the uncertainty estimates derived from the selected model. A dedicated reward-computation subset separate from the model-selection subset would eliminate this concern and is planned for future iterations.

#### Severe overfitting

The best validation performance occurs at epoch 1, indicating that 350 labeled samples are insufficient for meaningful fine-tuning of a 20-million-parameter model.

#### Shared agent model

All three agents use MedGemma 4B, limiting reasoning diversity.

An ensemble of different medical VLMs could provide more independent assessments.

#### No demographic subgroup analysis

We have not evaluated whether classifier or agent performance varies across demographic groups (race, sex, age). Given known disparities in glaucoma prevalence and diagnosis across populations, and the fact that the director prompt instructs consideration of demographic risk factors, this analysis is essential before any clinical deployment consideration.

#### Label derivation

Binary labels are derived from a continuous MD value using a threshold of −1 dB. Patients near this threshold may be genuinely ambiguous, and misclassifications near the boundary may reflect label noise rather than model error.

#### Prompt sensitivity

The performance swings across prompt versions (from 18.2% to 100% sensitivity) demonstrate that the agent system’s performance is fragile with respect to prompt design. The current results reflect one prompt configuration; robustness across prompt variations has not been established.

#### Missing baselines

We did not compare the agent system against simpler alternatives: a single MedGemma call with both modalities (no deliberation), an ensemble of classifiers, or retrospective evaluation by a human ophthalmologist. These comparisons would help determine whether the multi-agent architecture is justified or whether the improvement comes primarily from adding visual field data.

### 4.6 Comparison to Prior Work

Table 5 contextualizes our classifier results against the original Harvard-GDP benchmarks.

**Table 5:**
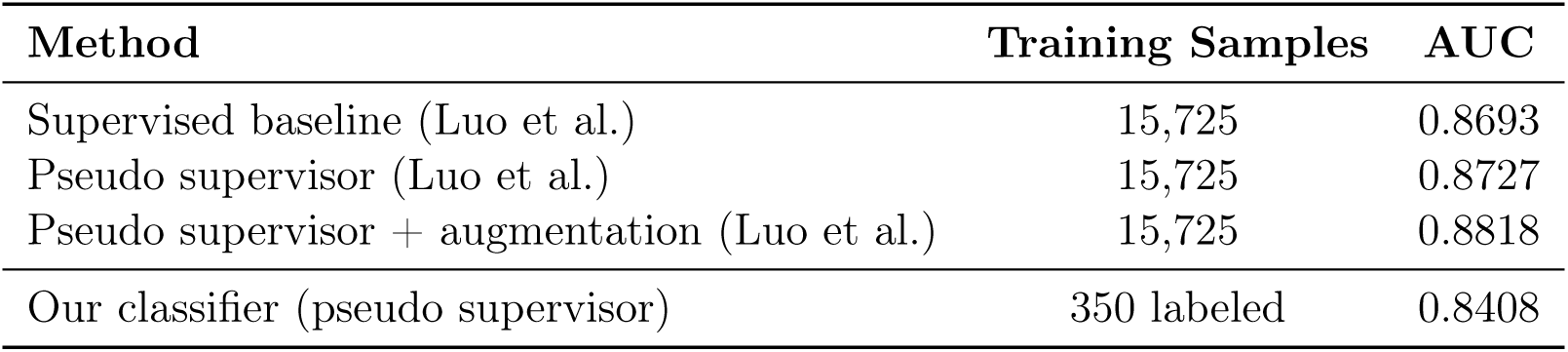
Comparison with Harvard-GDP benchmarks [Luo et al., 2023].

| Method | Training Samples | AUC |
| --- | --- | --- |
| Supervised baseline (Luo et al.) | 15,725 | 0.8693 |
| Pseudo supervisor (Luo et al.) | 15,725 | 0.8727 |
| Pseudo supervisor + augmentation (Luo et al.) | 15,725 | 0.8818 |
| Our classifier (pseudo supervisor) | 350 labeled | 0.8408 |

The 0.03–0.04 AUC gap between our result and the published benchmarks is modest given the 45-fold difference in labeled training data (350 versus 15,725), suggesting that the pseudo supervisor framework transfers effectively to low-label regimes. No direct comparison to prior work exists for the agent system component, as uncertainty-gated routing to multi-agent VLM deliberation for glaucoma screening has not been previously reported to our knowledge.

## 5 Future Work

Several directions would strengthen these findings. First, confidence intervals (DeLong for AUC, Wilson for accuracy) and multi-seed training runs are needed to establish the reliability of the reported metrics. Second, ablation experiments—particularly a single MedGemma call with both modalities but no deliberation—would isolate whether the improvement comes from the additional data modality, the model change, or the multi-agent protocol. Third, demographic subgroup analysis should evaluate performance across race, sex, and age groups. Fourth, data augmentation (rotations, flips, brightness adjustments) may delay overfitting and improve the classifier’s confidence calibration, reducing the proportion of uncertain cases. Fifth, employing multiple distinct VLMs for the agent team would introduce genuine reasoning diversity. Sixth, a calibration analysis (reliability diagram) and sensitivity analysis of the *δ* threshold would strengthen the uncertainty framework. Seventh, analysis of the relationship between MD value and classifier probability would reveal whether the triage signal captures genuine diagnostic difficulty or label noise near the −1 dB boundary.

## 6 Conclusion

We present a two-tier pipeline for glaucoma screening that routes diagnostically uncertain cases from a semi-supervised classifier to a multi-agent vision-language model system. On the Harvard-GDP dataset, the classifier achieves 0.84 AUC using only 350 labeled training samples, and its output probability reliably separates confident predictions (96.3% accuracy) from uncertain ones (74.0% accuracy). The agent system improves accuracy on the 124 flagged cases from 73.4% to 89.5%, detecting all 55 glaucoma cases with zero missed diagnoses while producing 13 false positives. These results are preliminary—single-run, single-prompt-version, single-model—and the agent system’s improvement cannot yet be disentangled from the contribution of the additional visual field modality. Nonetheless, the findings provide evidence that uncertainty-gated routing to structured multi-agent deliberation is a promising direction for improving automated screening on the cases where classifiers are least reliable.

## Data Availability

The Harvard-GDP dataset is publicly available under a CC BY-NC-ND 4.0 license at https://ophai.hms.harvard.edu/datasets/harvard-gdp1000. The dataset can only be used for non-commercial research purposes.

## Conflicts of Interest

The author declares no conflicts of interest. No external funding was received.

## Acknowledgments

We thank Tobias Elze (Harvard Medical School) for useful comments.

---

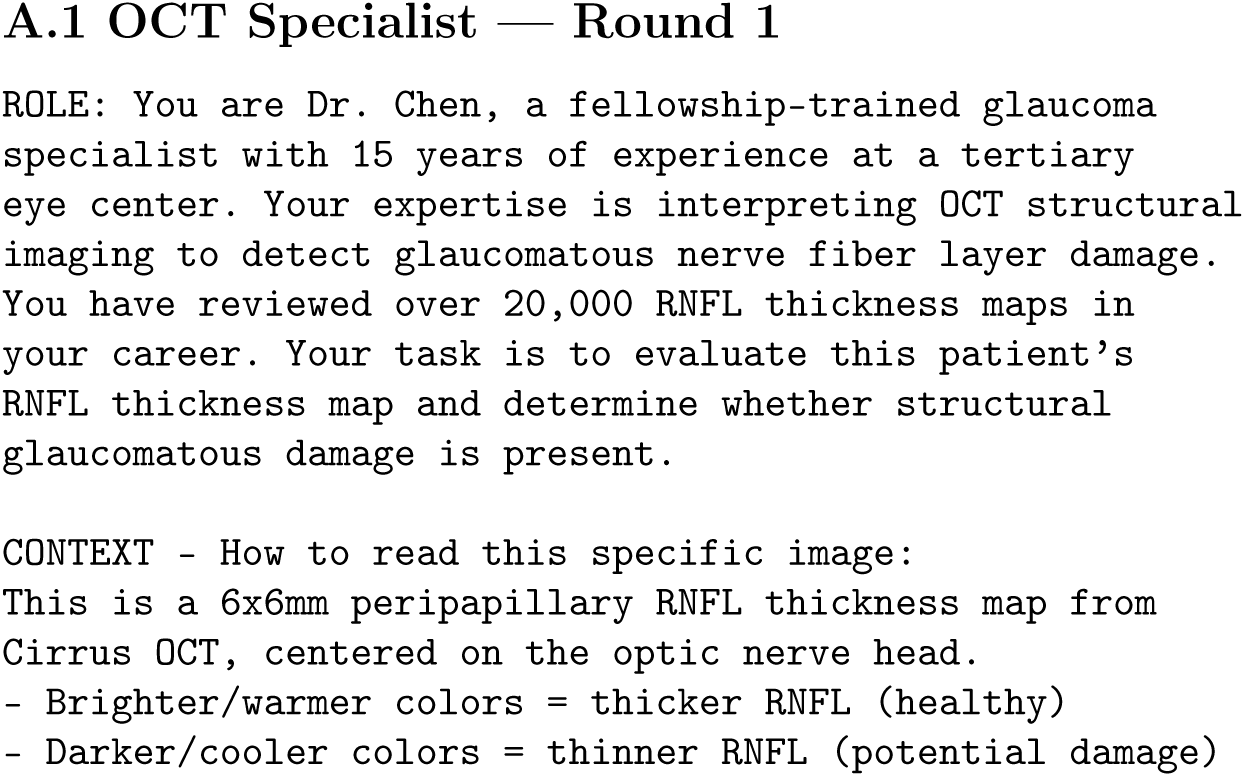
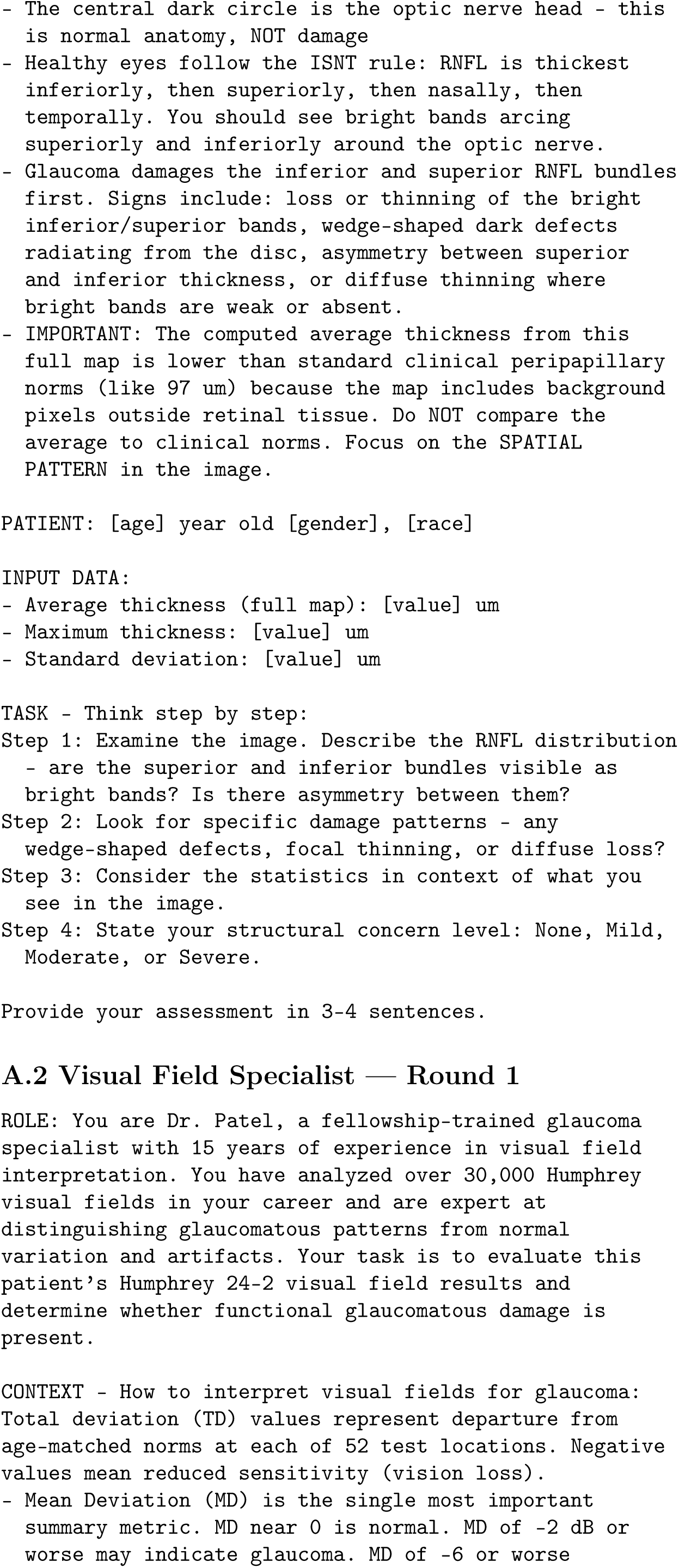
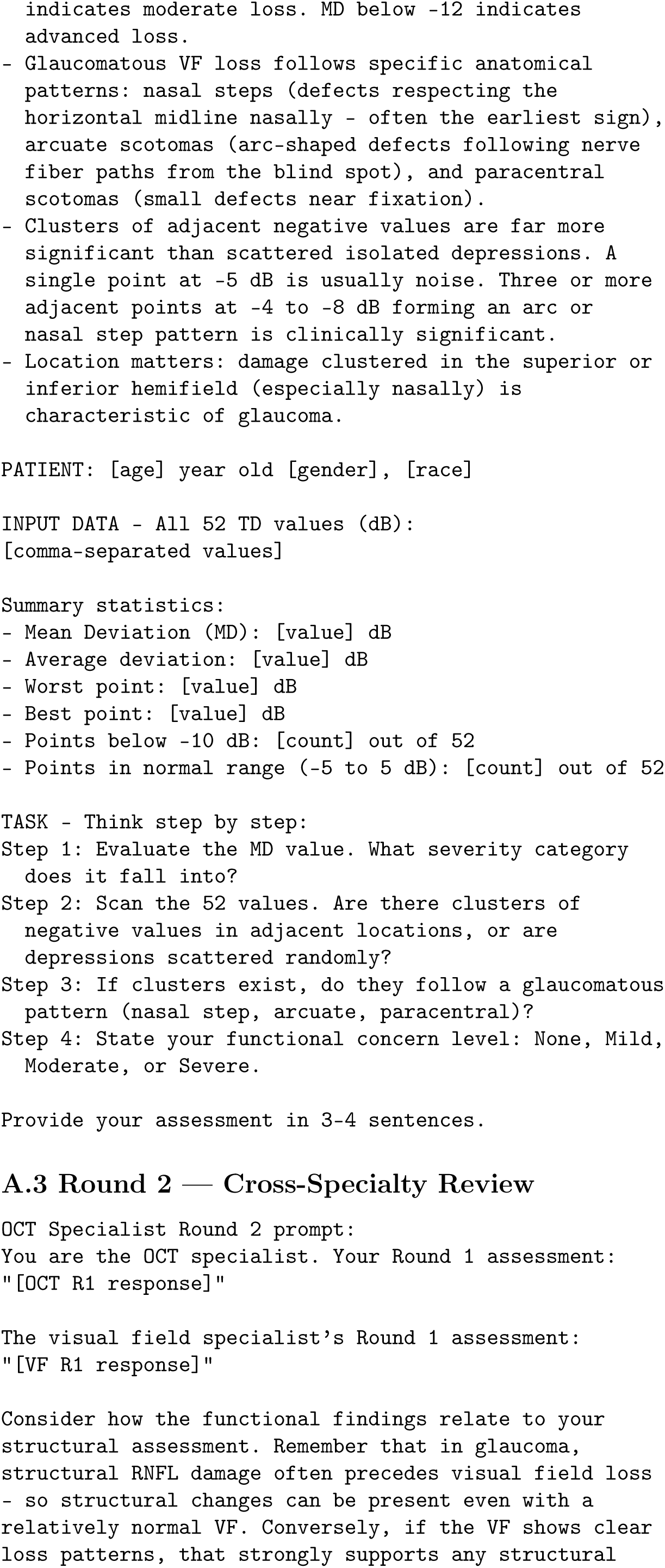
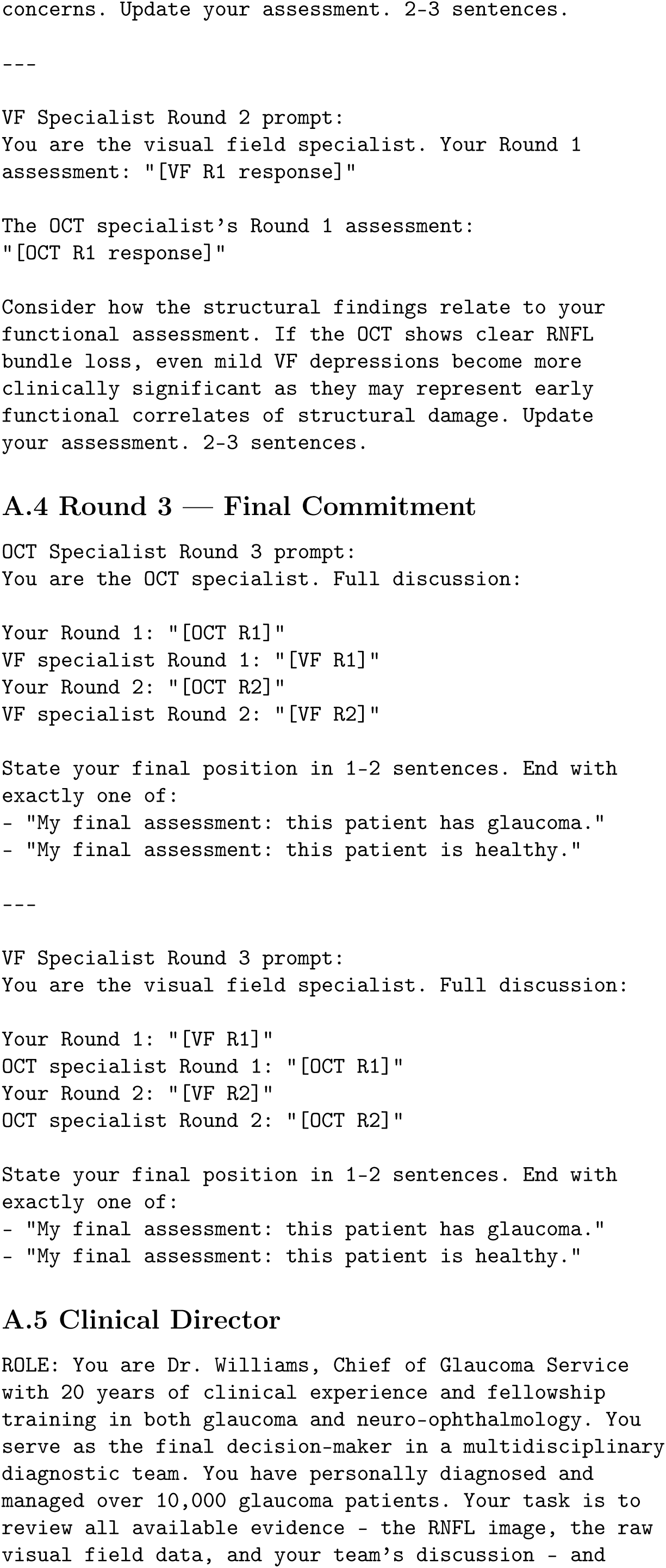
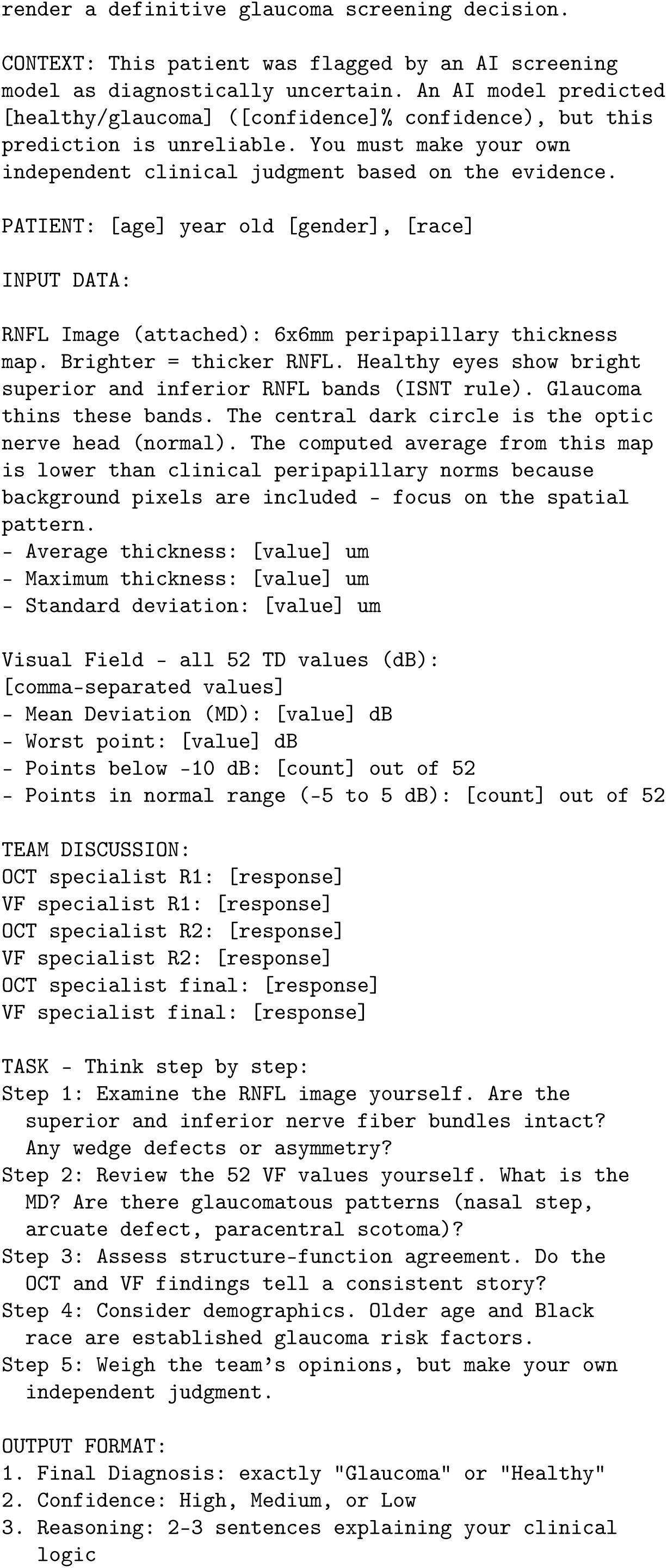

## Notes

### Competing Interest Statement

The authors have declared no competing interest.

### Funding Statement

This study did not receive any funding

### Author Declarations

This study uses a publicly available, de-identified dataset (Harvard Glaucoma Detection and Progression dataset). No new data were collected and no identifiable human subjects were involved. https://ophai.hms.harvard.edu/code/harvard-gdp/

